# Multi-omic analysis identifies a multi-step pathology in a case of multiple chorangioma syndrome in monochorionic twins

**DOI:** 10.1101/2024.12.03.24318397

**Authors:** Brandon M. Wilk, Manavalan Gajapathy, Donna M. Brown, Virginia E. Duncan, Elizabeth A. Worthey

## Abstract

**Background:** Chorangiomas, benign proliferative capillary lesions in the placenta, occur in approximately 1% of births, typically as a solitary nodule. In rare cases, multiple nodules develop, posing risks of fetal heart failure, hydrops fetalis, and intrauterine death due to altered placental hemodynamics. Although genetic and hypoxic factors have been hypothesized to drive aberrant angiogenesis, definitive evidence has been lacking. We report on a unique case of multiple chorangiomas in half of a shared placenta in monozygotic, monochorionic diamniotic (MCDA) twins, providing an unprecedented opportunity to explore impacts that molecular variation has on chorangioma formation.

**Results:** Whole genome and bulk RNA sequencing supported identification of early embryonic or germline and somatic variation. It revealed a likely pathogenic heterozygous frameshift deletion in EPAS1, a hypoxia-sensing transcription factor, with an early embryonic or germline origin. This variant likely impaired placental oxygen regulation and angiogenesis through its impact on VEGF-related pathways. Deleterious somatic mutations in COL1A1, FBXO11, and TRIM71 were observed within the chorangioma-affected tissue, along with increased expression of Leptin and DNA damage signatures consistent with oxidative stress. In contrast, the unaffected twin’s placental territory showed a different pattern of pathogenic somatic variation with the presence of a known pathogenic variant in MUTYH and signs of repair deficiencies. These findings highlight the presence of predisposing events and distinct molecular processes within each domain of the shared placenta. We propose that these molecular events, combined with environmental factors intensified by the MCDA pregnancy, likely contributed to chorangioma development..

**Conclusions:** Our study provides novel insights into the molecular basis of multiple chorangioma syndrome. To our knowledge, this is the first molecular evidence implicating both germline and somatic genetic involvement in this condition. The identification of molecular signatures previously associated with malignancy suggests that chorangiomas may share pathways with oncogenic processes. These findings highlight the importance of considering both genetic and environmental interactions in placental pathologies, offering potential implications for understanding and managing complex vascular and placental conditions, including preeclampsia, intrauterine growth restriction, and fetal vascular malperfusion.

## Background

Chorangiomas (chorioangiomas) are common non-cancerous vascular tumors in the placenta, occurring in about 1% of births, are generally present as solitary nodules, and are considered of no clinical significance[1, 2]. However, in rare situations where they become large (> 4cm in diameter) or are numerous, the risks of polyhydramnios, heart failure from arteriovenous shunting, growth restriction, hydrops fetalis, preterm delivery, sudden intrauterine fetal death, and/or stillbirth drastically increase as well as risks of preeclampsia and HELLP syndrome in the mother [1, 3, 4]. A recent review of multiple chorangioma cases found that only 4 of the 11 babies survived [3]. Chorangiomas are often identified in the second trimester or later [5], but they may also be discovered incidentally during other complications or after birth [6].

The etiology of chorangioma formation is not well understood [7, 8]. Chorangiomas and other hamartomas, including placental mesenchymal dysplasia, are noted in Beckwith-Wiedemann syndrome (BWS), though the genetic link is debated [7, 9–13]. A genetic predisposition has been suggested following a case of consecutive chorangiomas [14] and several cases of multiple chorangioma syndrome recurring across pregnancies [3]. However, reported cases of chorangioma formation with a possible genetic predisposition are quite rare [15].

Genetic studies have linked chorangioma formation to environmental factors including hypobaric hypoxia [13]. Chorangiomas are more prevalent in native high-altitude populations experiencing chronic hypobaric hypoxia [8, 16] and in less perfused areas of the placenta [9]. Vascular endothelial growth factor (VEGF) and abnormal angiogenesis in response to hypoxia have been implicated in chorangioma formation, as indicated by changes in the expression of growth, angiogenic, and anti-angiogenic factors in a case of recurrent multiple chorangiomas [17]. Increased rates of chorangioma formation have been noted in some studies of pregnancies of multiples [18, 19]. The molecular etiology of the association remains unclear [17, 18, 20].

We present a case of monozygotic, monochorionic diamniotic (MCDA) twins sharing a placenta, with extensive multiple chorangiomas confined to a single baby’s placental territory. The rarity of multiple chorangioma syndrome and the unique presentation in MCDA twins sharing a single placenta provided an ideal opportunity to investigate the molecular underpinnings of chorangioma formation. This case allowed us to examine whether specific molecular factors contributed to the differential placental outcomes. To explore why only one twin’s placental territory was affected, we conducted whole-genome and transcriptome sequencing of chorangioma and placental tissues, identifying distinct molecular variations, mutational signatures, and expression changes related to pathobiology. This case offers new insights into the molecular mechanisms of chorangioma development.

## Methods

### Sample preparation

Pathologic evaluation of the placenta was performed according to standard institutional protocols based on Amsterdam Criteria [21]. Formalin-fixed, paraffin-embedded (FFPE) blocks and H&E stained slides were prepared according to routine laboratory protocols. Chorangiomas were found to be extensive and confined to baby A’s placental territory (Figure 1). Regions of interest were identified on H&E stained slides and 2-mm core punches from unaffected villi, chorangioma tissue, and decidua from baby A’s placental territory and from normal villi from baby B’s placental territory were collected for sequencing (Figure 1). These were sent to the Vanderbilt University Medical Center (VUMC) VANTAGE lab for DNA and RNA isolation (the unaffected villi from baby A’s placental territory only had RNA isolated) and sequencing.

**Figure 1:**
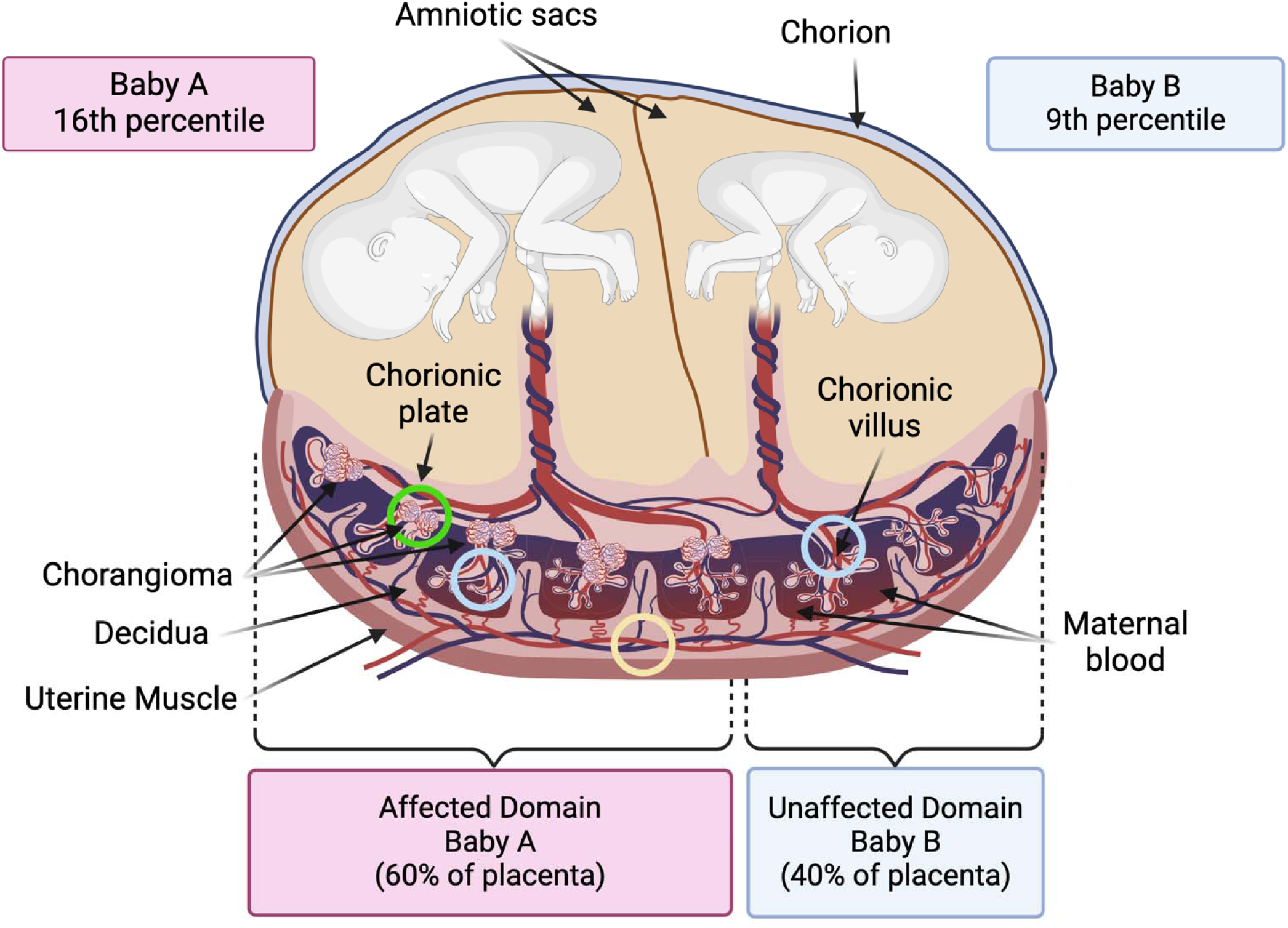
Twin and placental findings. Birth weight percentiles for each twin are shown. The placental chorionic vasculature (referred to as domains) is shown for both babies. Baby A’s domain contained an abundance of chorangiomas “Affected Domain”, whilst Baby B’s domain was free of chorangiomas “Unaffected Domain”. The location of the samples gathered for omic analyses are circled. Green denotes affected domain chorangioma tissue, light blue denotes unaffected villus tissue in each domain. Yellow denotes decidua tissue at the maternal surface.

### Whole Genome Sequencing and Analysis

DNA was isolated using Qiasymphony and/or FlexSTAR+ and quantified via Qubit and agarose gel electrophoresis to ensure sufficient yield and quality. Libraries were prepared with the Twist Biosciences kit (P/N: 104207) following manufacturer’s protocols. Library quality was assessed using the Agilent Bioanalyzer and quantified with qPCR using the KAPA Library Quantification Kit (P/N: KK4873) and QuantStudio 12K instrument. Library pools were combined in equimolar ratios, and cluster generation performed on a NovaSeq 6000 System. 150 bp paired-end sequencing targeted 400M reads per sample. Sequencing FASTQ files underwent QC, including read quality assessment, using Real Time Analysis Software and NovaSeq Control Software (v1.8.0), with MultiQC (v1.7) for data quality checks. Due to DNA degradation from fixation, each sample went through two library preparation and sequencing workflows to achieve sufficient mean coverage >30x.

Reads were downloaded to UAB’s Cheaha supercomputer and aligned to the GRCh38 reference genome using BWA-MEM (v0.7.15) [22]. Small, structural (SV), and copy number (CNV) variants were called followed GATK best practices [23] using haplotypecaller (v4.0.5.1) for germline variants, Mutect2 (v2.2) for somatic variants [24], and Manta (v1.6.0) for SVs and CNVs [25]. Somatic variants were called from chorangioma and villus tissue in tumor + normal mode, with the maternal surface (decidua) sample as the normal. Quality control (QC) for sequencing data, alignment, and variant calling was performed using QuaC (v1.0) [26]. All samples met threshold for analysis (mean coverage for chorangioma was 34x, normal villi 38x, and maternal surface 57x). Chorangioma and normal villi had a high degree of unalignable reads (36.4% and 23.5%).

FFPE fixation can induce DNA variation resulting in false variant calls [27, 28]. FFPE induced variants were identified and removed using FFPE somatic error detection methods as previously described [29] using the Ideafix detection tool [30]. Verification of FFPE-induced variant removal was performed using FFPESig downloaded on December 11th, 2023 [31]. Variant annotation, filtering, and analysis were performed using Codicem [32], VarSome [33], and cBioPortal [34–36] following best practices for variant analysis [37]. In brief, filtering was performed for population allele frequencies (e.g. gnomAD [38], *in silico* deleteriousness scores (e.g. CADD [39], PolyPhen-2 [40], and SIFT [41], and gene–phenotype associations relevant to the phenotype of interest (e.g. ClinVar[42] and Human Phenotype Ontology (HPO) [43]. Variant analysis was performed to identify germline/early placental variation and variation unique to the chorangioma. Germline variant pathogenicity was classified according to American College of Medical Genetics and Genomics (ACMG) guidelines [44] and somatic variant pathogenicity according to Association for Molecular Pathology (AMP) guidelines [45].

### Mutational Signature Profiling

To assess mutational processes in the chorangioma tissue, we profiled somatic variant calls from both chorangioma and normal villus tissue [46]. We used SigProfilerExtractor v1.1.21 [47] to extract and deconstruct the mutational signatures comparing them to the 96 reference Single base substitutions (SBS) contexts from COSMIC’s Mutational Signature catalog v3.3 [48]. The tool was run with default parameters, using GRCh38 as the reference genome, with minimum_signatures set to 1 and maximum_signatures set to 10.

### Chorangioma Subclone Analysis

Germline bi-allelic single nucleotide variants (SNVs) from WGS data were analyzed with CNVKit [49] for allele-aware CNV calling and copy number estimates. Somatic bi-allelic SNVs were filtered for a minimum coverage of 35x and variant allele fraction (VAF) greater than or equal to 0.08 and combined with genome-wide copy number estimates. These were then used for clone clustering and clonal population fraction estimation using PyClone-VI v0.1.3 [50]. PyClone-VI was run twice, using binomial and beta-binomial distributions, with the latter selected for further analysis based on recommendations for over-dispersed WGS data [51]. ClonEvol v0.99.11 [52] was used to predict clonal evolution based on PyClone-VI clustering. Settings for ClonEvol are detailed in the clonal-evo.Rmd document in the clonal evolution code repository (data and materials).

### RNA sequencing analysis

RNA was isolated from all tissue sites highlighted in Figure 1 and QC performed using Qubit or Picogreen with integrity assessed via BioAnalyzer or TapeStation. Library prep used NEBNext Ultra II Prep kits. Sequencing was conducted at VANTAGE on an Illumina NovaSeq 6000, generating paired end 150 bp reads, aiming for an average of 100 million reads per sample. Demultiplexed paired FASTQ outputs were returned. Initial QC was performed using FastQC, with adapter trimming and quality filtering via TrimGalore. QC metrics were poor due to RNA degradation, a known limitation with FFPE fixation [53, 54]. Given the extreme rarity of MCDA and multiple chorangioma events, advances in RNA sequencing from FFPE samples [55] and analysis improvements [56–58], we proceeded cautiously, making use of the nf-core RNA-Seq pipeline v3.6 [59] for alignment and expression quantification, with DESeq2 v1.28.0 for differential gene expression analysis.

## Results

### Case Description

MCDA twins were delivered without complications at 35 weeks via Cesarean section (C-section). Antenatal scans showed mild intrauterine growth restriction (IUGR) of Baby B (9th percentile), while the growth of baby A was at the 16th percentile. Although there were no signs of Twin-to-Twin Transfusion Syndrome (TTTS) C-section was performed due to nonreassuring fetal status in twin B. The babies both had Apgar scores of 8 and 9 and did well in the initial postnatal period. The mother had three prior pregnancies, including one loss for which we had no details. Pathologic examination of the shared placenta showed unequal placental territories (baby A: 60%, baby B: 40%). Overall placental weight was below the 5^th^ percentile for twin gestation (490g; 558-971g expected for 35 weeks). Multiple chorangiomas ranging in size from 0.1-1.5 cm in diameter were scattered diffusely throughout the placental parenchyma, entirely confined to the placental territory of baby A (Figure 1). WGS confirmed the twins were monozygotic, originating from a single genome.

### Germline variation in chorangioma formation

SV and CNV analysis revealed no variants of note, consistent with literature reports [20]. A novel likely pathogenic heterozygous near-splice 5-base frameshift deletion (c.22_26del, p.Lys8GlufsTer2) in Endothelial PAS Domain Protein 1 (EPAS1) was identified in the chorangioma tissue, decidua from baby A’s placental territory, and normal villi from baby B’s placental territory, indicating an early embryonic or germline origin (Table 1). RNA-Seq data confirmed expression of both wildtype and frameshift *EPAS1* alleles (Supplemental Figure 1).

**Table 1:** Somatic and germline variants of interest. Classification of variants of interest based on ACMG and AMP guidelines. * VAF provided for EPAS1 within Chorangiomas, normal villi from baby B’s placental territory, and decidua. Germline and somatic variation was identified with somatic variation differing between affected and unaffected samples.

| Tissue | Allele Origin | Gene | Variant | Classification | VAF* | GnomAD AF |
| --- | --- | --- | --- | --- | --- | --- |
| All | Germline or Early Embryonic | EPAS1 | NM_001430.5: c.22_26del (p.K8Efs*2) | Likely Pathogenic | 51/52 /39 | Not seen |
| Unaffected Villi | Somatic | MUTYH | NM_001048174.2: c.650G>A (p.R217H) | Pathogenic/ Tier II | 7.4 | 0.000024 |
| Chorangioma | Somatic | COL1A1 | NM_000088.4: c.1588G>A (p.G530S) | Pathogenic/ Tier II | 8.3 | Not seen |
| Chorangioma | Somatic | FBXO11 | NM_001190274.2: c.856G>T (p.G286*) | Likely Pathogenic/ Tier II | 8.6 | Not seen |
| Chorangioma | Somatic | TRIM71 | NM_001039111.3: c.2386C>T (p.R796C) | Likely Pathogenic/ Tier II | 11 | Not seen |

*EPAS1*, also known as Hypoxia-Inducible Factor 2 Alpha (HIF2A), is highly expressed in lung, adipose, and placental tissues, playing a central role in hypoxia response by regulating genes involved in metabolism, proliferation, angiogenesis, erythropoiesis, and VEGF expression [60–62]. *EPAS1* plays a crucial role in placental structure, angiogenesis, and embryonic development, supporting blood vessel and lung tubular system formation, particularly under hypoxic conditions [63–66]. Germline gain-of-function variants in *EPAS1* cause dominant familial erythrocytosis 4, marked by erythrocytosis and deep venous thrombosis [67]. Germline, somatic, or postzygotic gain-of-function *EPAS1* variants in early embryogenesis are also associated with congenital polycythemia, somatostatinoma, Pheochromocytomas and Paragangliomas Syndrome, and Pacak–Zhuang syndrome [68–71]. Altered *EPAS1* expression has been linked to PI3K/mTORC2 activity, tumor angiogenesis, and malignant tumor progression [72–75]. In Tibetan populations, *EPAS1* variants reduce expression in placental tissue under high-altitude hypoxia, blunting the physiological response to chronic hypoxia and coinciding with an increased incidence of chorangioma formation [16, 76–80].

Homozygous EPAS1 knockout in mice is lethal post-vasculogenesis due to improper blood vessel fusion in the yolk sac and embryo, and failure to form larger vessels [81, 82]. Heterozygous mice are viable but show blunted responses to chronic hypoxia [81, 83]. EPAS1 also directly regulates DNMT1 in lung tissue, linking it to DNA methylation defects and imprinting issues associated with about 50% of Beckwith-Wiedemann syndrome cases [84, 85].

### Somatic variation

No somatic CNVs or SVs of interest were identified. A pathogenic somatic variant (c.650G>A, p.R217H) in *MUTYH* was identified in normal villi (Table 1). Its expression couldn’t be confirmed in RNASeq data due to lack of coverage. This gene encodes a glycosylase involved in base excision repair (BER) of oxidative DNA damage, specifically removing adenine mispaired with 8-oxoguanine [86, 87]. The variant lies adjacent to the endonuclease active site, essential for repair initiation. Found constitutively, this variant causes familial adenomatous polyposis and other cancer syndromes [86, 88] and has also been identified as a somatic variant in malignant tumors [89].

Within the chorangioma, three somatic variants of interest were identified (Table 1), though their expression couldn’t be confirmed in the RNASeq data due to insufficient gene coverage or absence of the allele. One pathogenic variant (c.1588G>A, p.G530S) in *COL1A1* encodes the alpha-1 chain of type I collagen, essential for tensile strength and stability in connective tissues. Deleterious germline variants in *COL1A1* are associated with osteogenesis imperfecta and Ehlers-Danlos syndrome [90–92]. Somatic variants in *COL1A1* influence tumor extracellular matrix, affecting tissue stiffness and tumor progression [93]. In the placenta, *COL1A1* is essential for structural integrity, guiding cellular behaviors, activating PI3K-AKT signaling, epithelial-to-mesenchymal transition, angiogenesis, and hypoxia response [94, 95]. Disruptions in these processes can lead to vascular and placental abnormalities [96–98].

A likely pathogenic truncating variant (c.856G>T, p.G286Ter) was identified in *FBXO11*, an E3 ubiquitin ligase essential for tagging proteins for degradation to maintain cellular protein balance [99–101]. Dysregulation of *FBXO11* is linked to cancer and developmental disorders and is often inactivated in lymphoma and other tumors, contributing to abnormal growth and tumor development [102, 103].

A second likely pathogenic missense variant (c.2386C>T, p.R796C) was identified in *TRIM71*, an E3 ubiquitin ligase that regulates the cell cycle via RNA binding to the 3’ UTR of *CDKN1A/p21*, promoting NMD and embryonic stem cell proliferation [104, 105]. *TRIM71* is highly expressed in extravillous trophoblasts (EVTs) in the placenta [106–108]. Germline variants in *TRIM71* are associated with autosomal dominant congenital hydrocephalus, characterized by cerebral ventriculomegaly, and altered expression has been linked to tumorigenesis [105, 109–111].

### Differential expression

Differential expression analysis showed a 3.1-fold increase (p adj. < 0.05) in Leptin (*LEP*) expression in chorangioma tissue compared to normal villous tissue. Leptin, a multifunctional endocrine protein, regulates angiogenesis, PI3K-AKT, and VEGF pathways in various cell types, including cancers [112]. A rare somatic SNV (chr7:g.128236766A>T) in an enhancer of *LEP* was found in the chorangioma tissue, though its effect is unclear. No significant expression differences were observed for genes with WGS-identified variants (Table 1).

### Chorangioma clonal evolution

We hypothesized that somatic variation patterns in these chorangiomas might resemble those in cancer, where subclonal architecture could reveal chorangioma-specific mutational processes. Clonal analysis identified a linear model of molecular evolution, showing selective pressure favoring expansion of a subclonal population (population 3) exclusively in the chorangioma (Figure 2). This supports the idea that mutational processes impact these tissues before or during chorangioma development.

**Figure 2:**
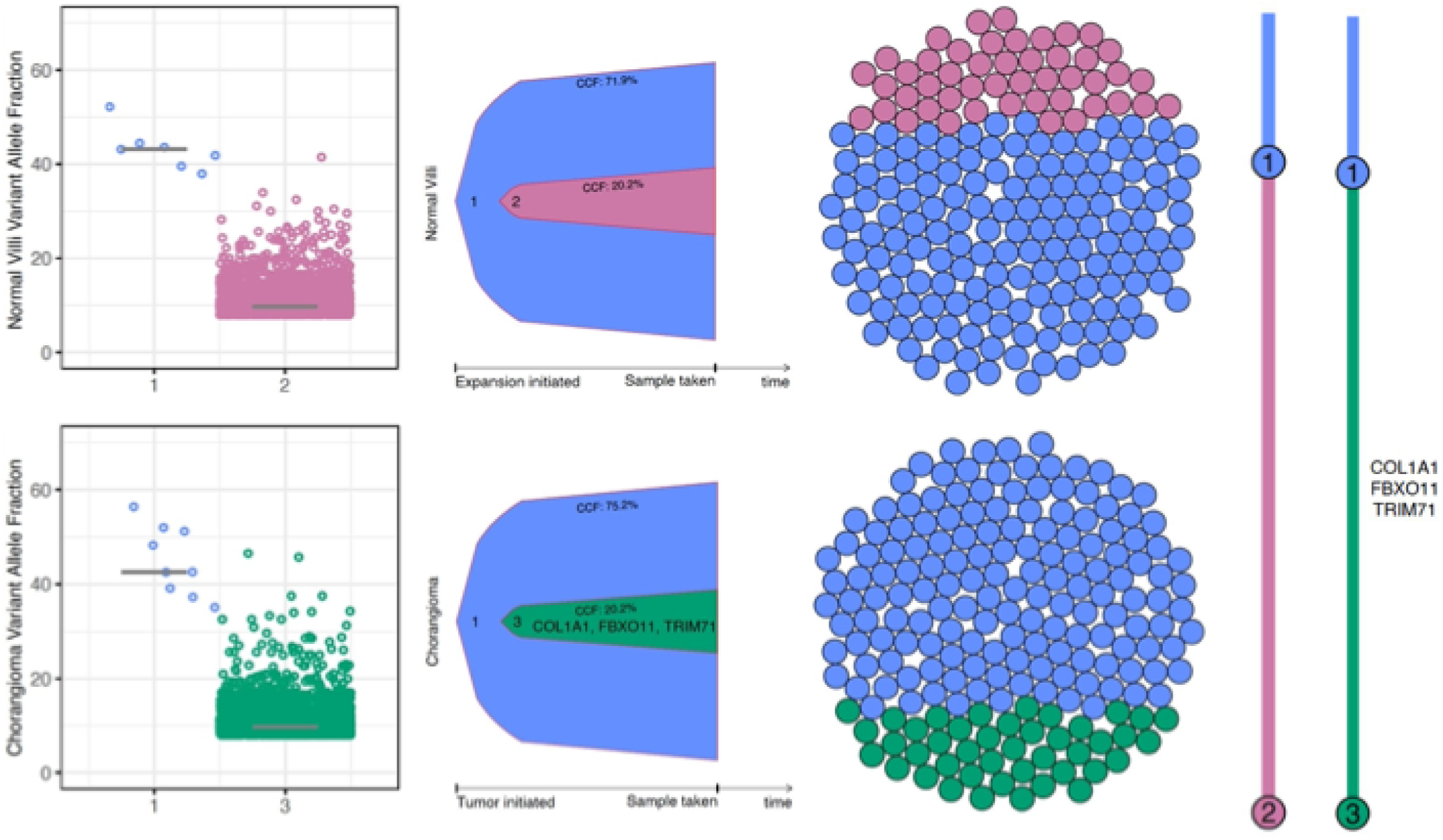
Chorangioma clonal evolution. Scatter plots show the cancer cell fraction (CCF) of each clonal cluster. Bell plots represent clonal dynamics over time and the sphere of cells depict the estimated subpopulation fractions. The branching clonal evolutionary model (far right) shows the predicted clonal evolution of chorangioma tissue and normal villi. It is clear from this model that cluster 3 indicates a subclonal population in the chorangioma not seen in the normal villi.

### Mutational signatures of chorangiomas

Somatic variation can result from events that leave distinct mutational signatures reflecting underlying processes, including replication errors, repair deficiencies, environmental exposures, and oxidative stress. The dominant signature in baby A’s chorangioma sample, SBS18 (Figure 3, Supplemental Figure 2), is associated with DNA damage from reactive oxygen species (ROS)[113, 114]. This may be indicative of elevated oxidative stress due to rapid cell proliferation, high metabolic activity, or abnormal vasculature with frequent hypoxia-reperfusion cycles not present in normal placenta [115–117]. SBS18 has been observed at lower levels in healthy placental tissue [46]. SBS51, also present, is linked to sequencing artifacts [48, 118].

**Figure 3:**
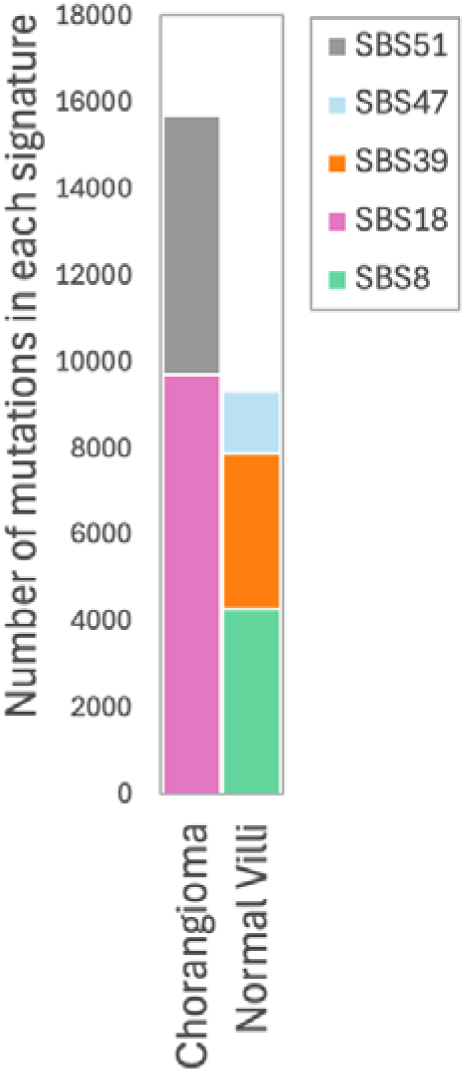
Mutational signature analysis. The majority of somatic variation identified in the chorangioma sample was attributed to the ROS induced SBS18 signature. An equal amount of variation in the normal villi was attributed to SBS8 and SBS39 signatures. The chorangioma and to a lesser degree the normal villi sample also had patterns of somatic variants attributable to likely sequencing artifacts (SBS47 and SBS51).

SBS8 is the dominant mutational signature in the normal villous tissue from baby B’s placental territory, associated with deficiencies in homologous recombination/nucleotide excision repair (HR/NER)[119, 120] and previously unreported in placental tissue. The placenta’s rapid growth and physiological stress may increase DNA damage, activating alternative repair pathways. If so, SBS8 likely reflects high demand on repair systems during normal development rather than pathology. SBS39 has been linked to DNA polymerase eta (POLH) A>T mutations during DNA replication under UV exposure [121], and noted in embryonal rhabdomyosarcoma, but the association here is unclear. SBS47, a minor component associated with sequencing artifacts [122], is smaller in normal villi than in chorangioma tissue, likely due to differences in DNA quality between tissues.

## Discussion

Placental development is a carefully coordinated process influenced by oxygen levels, which play a crucial role in placentogenesis by regulating villous vascularization, and trophoblast differentiation and proliferation. Chronic environmental hypoxia or genetic changes are hypothesized to drive aberrant angiogenesis and chorangioma formation, though definitive evidence has been limited. Our findings from a case of multiple chorangiomas in half of a shared placenta of monozygotic MCDA twins suggest a multi-step pathology leading to chorangioma development (Figure 4).

**Figure 4:**
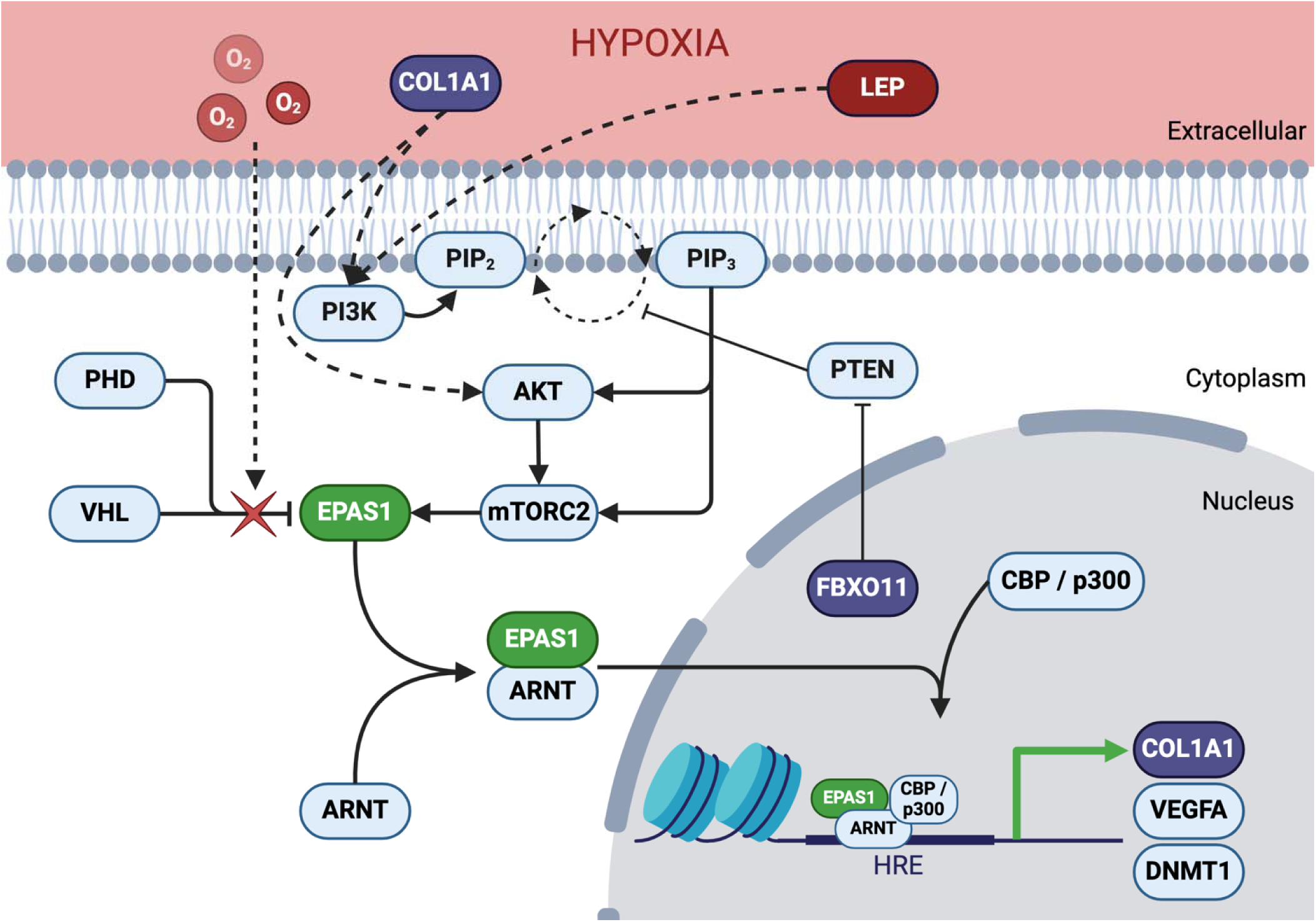
Hypoxia driven EPAS1 Stabilization. Under normoxia, PHD hydroxylates EPAS1 (in green) allowing VHL to ubiquitinate EPAS1 leading to proteasomal degradation. Under hypoxia, PHD fails to hydroxylate EPAS, preventing VHL binding, leading to EPAS1 accumulation. This triggers EPAS1 dimerization with ARNT, nuclear translocation, recruitment of CBP/p300, and activation of target genes containing hypoxia response elements in their promoter. Accumulation of ROS increases somatic mutation in genes within the chorangioma (in purple). These ROS driven alterations are identified as ROS damage and deficient repair mutational signatures and contribute to abnormal capillary proliferation and genomic instability, leading to altered placental growth dynamics. Changes in expression (in red) also impact angiogenesis, vascularization, and placental integrity. We hypothesis that combination of EPAS1 variation, MCDA twin pregnancy, and COL1A1, FBXO11, and TRIM71 somatic mutation in the baby A’s larger placental territory impacted placental development, blood vessel formation and tissue integrity, and angiogenesis, ultimately resulting in the abnormal vascular structures of the chorangiomas in that share only.

We identified a heterozygous frameshift deletion in *EPAS1* across all sequenced tissues, suggesting an early embryonic or germline origin in pre-morula development (days 1-3). We hypothesize this variant likely impairs the placenta’s ability to sense and adapt to the low-oxygen environment of early pregnancy, where *EPAS1* typically stabilizes to support development. Here, *EPAS1* haploinsufficiency likely blunted this adaptation, and the pregnancy’s development into MCDA twins with a shared placenta (days 4–8) further compromised the placenta’s capacity to respond to maternal oxygen variations. In MCDA pregnancies, high resource demands and limited adaptive capacity increase stress and hypoxia, making placental tissue more susceptible to oxidative damage. We suggest that early first-trimester oxidative stress from MCDA and *EPAS1* alteration in this case likely raised reactive oxygen species, leading to DNA damage.

In baby A’s placental territory, we identified deleterious somatic mutations in *TRIM71*, *FBXO11*, and *COL1A1*, which likely intensified ROS-related damage, promoted abnormal angiogenesis, and altered hypoxic responses, consistent with observed mutational signatures. We propose that these variants may have driven localized angiogenesis and vascular proliferation, disrupting hypoxic signaling and contributing to chorangioma development. While our findings underscore the genetic factors in chorangioma formation, non-genetic factors, such as asymmetrical placental territory sizes, maternal health, or in utero hypoxic events, would also be expected to have played a role.

While our findings are novel, limitations exist. RNA degradation from FFPE limited expression analysis, and DNA degradation may have affected mutational signature profiling [31]. Analysis from fresh or frozen tissues would be beneficial. Although we used an optimized pipeline for clonal evolution [123], identifying all subclonal populations is limited by standard WGS sequencing depth and single tissue sampling [124]; greater coverage would have benefited analyses. This study provides a detailed molecular analysis of an ultra rare and unusual case of multiple chorangioma syndrome in MCDA twins. Statistical interpretation in single-case studies remains challenging, as rare variants in low-prevalence conditions rarely meet traditional significance thresholds. Additional research is needed to determine if *EPAS1* involvement is specific to this case or indicative of broader patterns in multiple or large chorangiomas. The rarity of multiple chorangiomas will make gathering additional samples a lengthy process. We therefore interpret our findings with caution, acknowledging that the observed *EPAS1* variant and molecular signatures may represent just one of multiple possible pathways to chorangioma formation.

In the meantime, modeling these alterations in systems that closely resemble human placental physiology, such as guinea pigs [125] or baboons [126], would enhance understanding of *EPAS1* haploinsufficiency and hypoxia-related effects. Applying single-cell and spatial approaches would also improve resolution at the maternal placental surfaces and support deeper understanding of clonal complexity. Additionally, follow-up studies using CRISPR/Cas9 gene editing in trophoblast or endothelial cell lines would help assess impacts on hypoxia response, angiogenesis, and DNA repair pathways. Together, these functional studies would confirm the role of these variants in chorangioma formation and expand our understanding of placental pathophysiology.

In summary, we present molecular findings from a unique case of multiple chorangioma syndrome in monozygotic MCDA twins, affecting only one twin’s placental territory. To our knowledge, this is the first report to implicate a molecular mechanism in multiple chorangioma syndrome and the first to propose the interaction of germline and somatic variants in the pathobiology of these tumors. While chorangiomas are typically benign, our analysis suggests that mutational processes with potential oncogenic activity beyond abnormal angiogenesis can be involved. This case underscores the complex genetic and environmental interplay in chorangioma formation, advancing our understanding of placental biology and the role of genetic-somatic interactions in vascular syndromes. These insights lay a foundation for future research, highlighting specific genetic and environmental factors that warrant investigation as similar cases emerge.

## List of abbreviations

ACMG: American College of Medical Genetics and Genomics
AMP: Association for Molecular Pathology
ARNT: Aryl Hydrocarbon Receptor Nuclear Translocator
BER: base excision repair
BWS: Beckwith-Wiedemann syndrome
CBP: CREB-binding protein
CCF: cancer cell fraction
CDKN1A/p21: Cyclin Dependent Kinase Inhibitor 1A
COL1A1: collagen type I alpha 1 chain
COSMIC: Catalogue Of Somatic Mutations In Cancer
CREBBP: CREB-binding protein
DNMT1: DNA methyltransferase 1
DTL: denticleless E3 ubiquitin protein ligase homolog
ECM: extracellular matrix
EP300: E1A binding protein p300
EPAS1: endothelial PAS domain protein 1
FBXO11: F-box protein 11
FFPE: Formalin-Fixed Paraffin-Embedded
HELLP: Hemolysis, Elevated Liver enzymes and Low Platelets
HIF1A: hypoxia inducible factor 1 subunit alpha
HIF1B: hypoxia inducible factor 1 subunit beta
HIF2A: hypoxia inducible factor 2 subunit alpha
HIF3A: hypoxia inducible factor 3 subunit alpha
HRE: hypoxia response element
IUGR: intrauterine growth restriction
LEP: leptin
MCDA: Monochorionic, diamniotic
MUTYH: mutY DNA glycosylase
NMD: nonsense-mediated mRNA decay
OMIM: Online Mendelian Inheritance in Man
PHD: hypoxia-inducible factor prolyl hydroxylase
PI3K: Phosphoinositide 3-kinases
PI3K-AKT: Phosphoinositide 3-kinases-Akt serine/threonine kinase pathway
POLH: DNA polymerase eta
PTEN: phosphatase and tensin homolog
RNA-Seq: Bulk RNA sequencing
ROS: reactive oxygen species
SBS: single base substitution
SNV: single nucleotide variant
TRIM71: tripartite motif containing 71
VAF: variant allele fraction
VEGF: vascular endothelial growth factor
VHL: von Hippel-Lindau tumor suppressor
VUS: variant of unknown significance
WGS: whole genome sequencing

## Declarations

### Ethics approval and consent to participate

All research conducted during this study has been completed in accordance with guidelines from the Declaration of Helsinki and IRB approved under a category 4 exemption. IRB review was conducted by the University of Alabama at Birmingham Institutional Review Board under reference IRB-300004310.

### Consent for publication

Not applicable

### Availability of data & materials

Somatic variants, code, and documentation on the mutational signature analysis can be found in our GitHub repository at https://github.com/uab-cgds-worthey/somatic-mutation-signal-identification. The input somatic variants, CNVKit genome-wide copy-number estimates, input merging and running instructions for variant clustering using PyClone-VI and clonal evolution by ClonEvol are available in our GitHub repository at https://github.com/uab-cgds-worthey/multiple-chorangiomas-subclone-analysis. The principal author takes full responsibility for the data presented in this study, analysis of the data, conclusions, and conduct of the research. Datasets generated from the analysis presented will be available from the corresponding author on reasonable request.

### Authors’ contributions

EAW obtained funding and supervised the study. VD performed the pathology work up, tissue collection, fixation, storage, physical tissue processing and send out for sequencing. VD also obtained IRB exemption for conducting this research. BMW and EAW set up the experimental design and hypothesis generation. BMW performed data processing and analysis. MG performed structural and copy number variant analysis. BMW and EAW performed variant analysis. BMW performed RNA-Seq analysis. All authors participated in review of results. BMW and EAW wrote the initial drafts of the manuscript. All authors read and refined subsequent drafts and approved the final manuscript.

## Acknowledgements

This work was supported by UAB Start-Up funds (to EAW). We would like to acknowledge the support of the Vanderbilt VANTAGE sequencing lab for DNA and RNA sequencing. We thank Lara Ianov, Ph.D. and Elizabeth Wilk, M.Sc. for their insight and assistance with RNASeq analysis.

---

**Supplemental Figure 1:**
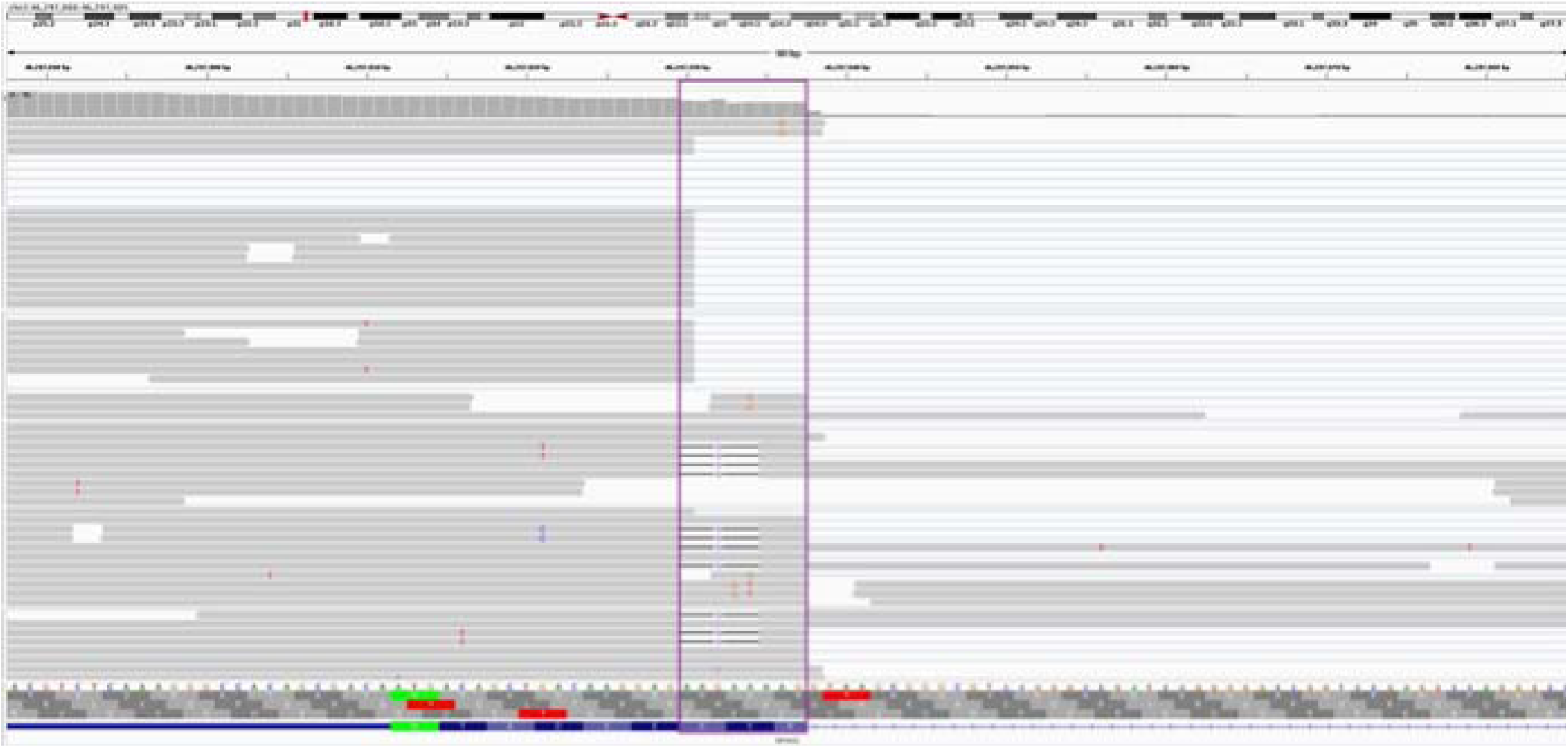
*EPAS1* variant and wildtype allele expression. A representative alignment of RNASeq reads highlighting the 5bp frameshift (within the purple highlighted box) identified in EPAS1 does not alter splicing. Some reads containing the 5bp deletion are aligned such that the ‘AAG’ before the splice site is aligned here (reads in lower half of alignment). The repetitive ‘AAG’ sequence here and at the 3’ splice junction (not depicted) result in a situation where the first ‘A’ base is aligned here and the subsequent ‘AG’ is aligned at the 3’ splice junction with a single gap (reads in the upper half).

**Supplemental Figure 2:**
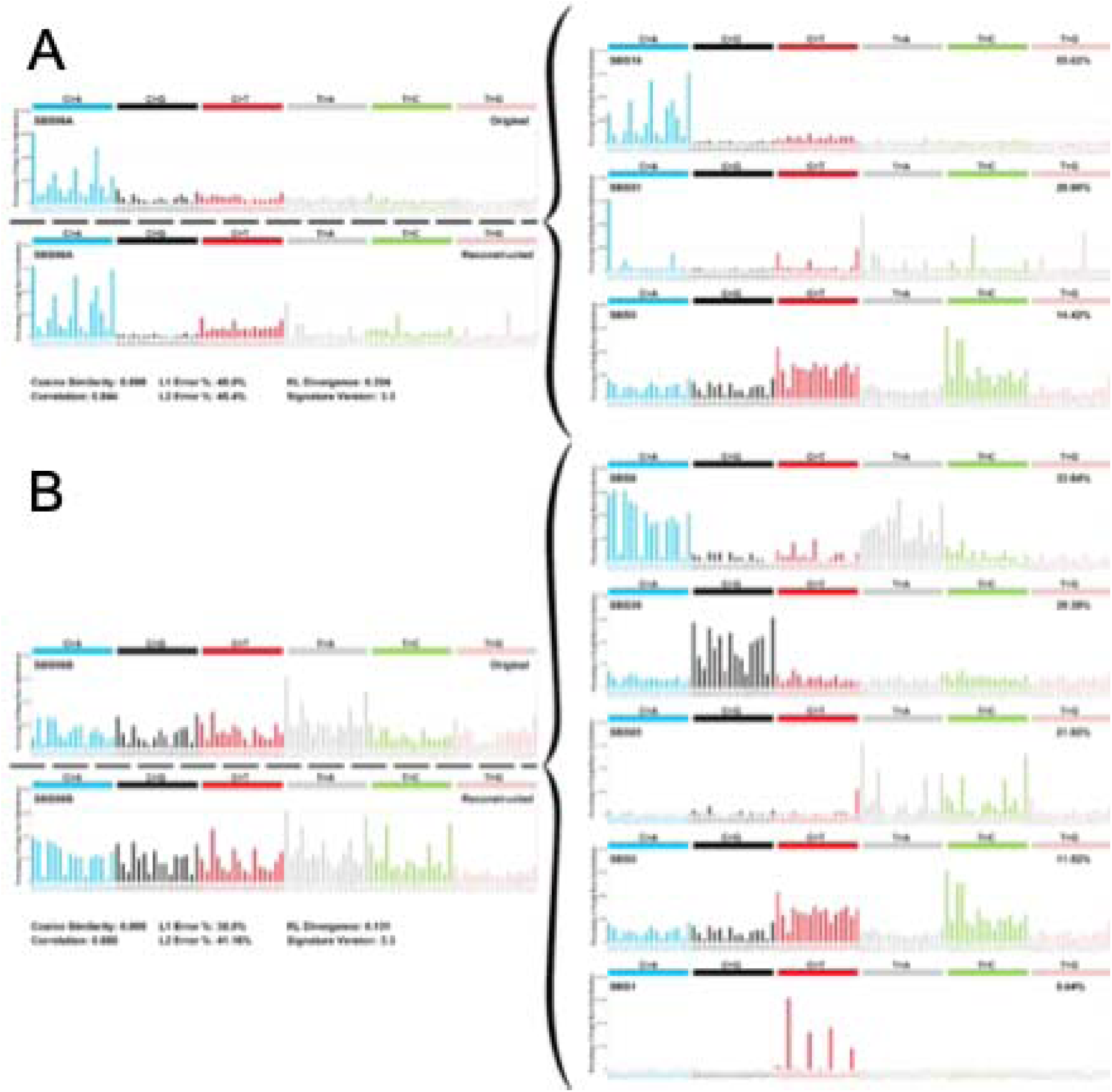
Single base substitution (SBS) mutational signatures in chorangioma tissue and unaffected villi. Different composites of the 96 COSMIC SBS reference signatures were identified in both tissues. Panel A shows the identification of mutational signatures in chorangioma tissue and Panel B for the unaffected villi. Each plot shows the identified signature and reconstruction from estimated proportions of the 96 COSMIC SBS reference signatures (right side plots in each panel).

